# Language and Social Regions Are Affected in Toddlers with Autism and Predict Later Language Outcome

**DOI:** 10.1101/2022.10.25.22281531

**Authors:** Kuaikuai Duan, Lisa Eyler, Karen Pierce, Michael V. Lombardo, Michael Datko, Donald Hagler, Vani Taluja, Javad Zahiri, Kathleen Campbell, Cynthia Carter Barnes, Steven Arias, Srinivasa Nalabolu, Jaden Troxel, Anders M. Dale, Eric Courchesne

## Abstract

Autism spectrum disorder is a heterogeneous neurodevelopmental disorder. Early brain overgrowth yet reduced cerebellar size is well recognized for autism, but cortical regions involved show inconsistent patterns of alteration. No complete and replicable map of early regional brain size alterations has been charted. It is also not clear whether individual differences in brain size relate to autism symptom severity and cognitive deficits and predict later language outcomes. We leveraged structural MRI data from 166 autistic and 109 typical developing toddlers to comprehensively and systematically investigate regional gray matter volume alterations and cortical surface area and thickness perturbations in autism compared to typical developing toddlers using linear mixed-effect models. We then examined their replicability in an independent cohort of 38 autistic and 37 typical developing toddlers. We further investigated associations between regional brain size and symptom severity, Mullen and Vineland cognitive performance using linear regression models. Lastly, we investigated whether early brain size (at intake mean age of 2.5 years) can improve support vector machine prediction of language outcome at 3-4 years of age when added to a model containing intake clinical and behavioral measures. Compared to typical developing toddlers, autistic toddlers presented larger or thicker lateral temporal regions, smaller or thinner frontal lobe and midline structures, larger callosal subregion volume, and smaller cerebellum. Most of these differences were replicated in an independent toddler cohort. Moreover, the identified gray matter alterations were related to autism symptom severity and cognitive impairments at intake, and, remarkably, they improved the accuracy for predicting later language outcome beyond intake clinical and demographic variables. Gray matter volume, thickness, and surface area in regions involved in language, social, and face processing were altered in autistic toddlers. Alterations in these regions are major early-age developmental attributes of autism. The early-age alterations in these cortical attributes in different regions may be the result of dysregulation in multiple neural processes and stages, consistent with prenatal multi-process, multi-stage models of autism. Here we also show these gray matter alterations are promising prognostic biomarkers for language outcome prediction.

## Introduction

Autism spectrum disorder (ASD) is a neurodevelopmental disorder characterized by social and communicative deficits and repetitive behaviors emerging at 1-4 years old ^1,2^. ASD affects approximately 1 in 54 children in the United States ^3^. The high prevalence rate of ASD and associated social and language deficits significantly elevate the risk of adverse outcomes for individuals with ASD and increase the burden for the involved families and the whole society. Clinical heterogeneity of ASD is considerable ^4-8^: Some toddlers benefit from contemporary ABA treatments, but others do not. Some toddlers may earn a college degree and live independently, but others remain minimally-verbal with life-long struggles with social communication. While language and social symptoms improve with age in some toddlers, they do not for others, and such outcome differences are not clearly predictable from clinical scores at very early ages ^4-8^. Characterizing ASD neuropathology at the age of clinical onset, and how it relates to clinical heterogeneity, is essential for aiding early diagnosis, prognosis, and early interventions.

Converging evidence from neuroanatomical studies suggests brain overgrowth in young kids with ASD ^9-15^, especially in frontal and temporal regions ^2,14-17^, while other brain regions showed inconsistent brain alteration patterns in ASD. For example, both volume increases and reductions have been reported in the amygdala ^18-20^, corpus callosum ^21-24^, and cerebellum ^25-28^. The inconsistent results may be due to cohort (e.g., subject characteristics), MRI scanner, preprocessing pipeline, and analytical methodology differences ^20,29^. Moreover, most studies focused on global measures or single regions (e.g., amygdala, cerebellum, and corpus callosum) and single morphometries (e.g., volume, surface area, cortical thickness) of interest that may be relevant to ASD. In the cortex, surface area and cortical thickness are dissociable features ^30^; examining potential alterations in both features in the same sample may point to distinct biological origins of cortical gray matter changes. No study of brain alterations in young kids with ASD has yet examined regional differences across the brain and examined volume, cortical thickness, and surface area in a comprehensive manner.

Brain size alterations have been widely reported to underlie language and social deficits and facial recognition impairment in ASD. For example, volumes in frontal and temporal regions were related to repetitive behavior and social and communication deficits in ASD as revealed in an unbiased voxel-based morphometry study ^25^ or a source-based morphometry (a multivariate approach) study ^31^. Moreover, Dziobek and colleagues identified that increased cortical thickness in the fusiform gyrus was associated with more severe face processing impairments in individuals with autism ^32^. These studies used a cross-sectional design and examined an older sample among whom compensatory neural alterations may have resulted from behavioral challenges rather than caused them.

There are heterogeneous developmental courses in ASD; some ASD toddlers get better, and others get worse with age ^33-35^. Our previous work demonstrated that degree of functional hypoactivation of ASD toddlers in the temporal region in response to a language task markedly improved the accuracy for classifying language outcome when combined with behavioral and clinical variables ^33^. However, it’s not clear whether structural alterations of subcortical and cortical regional size identified at the earliest clinic visit contribute to discriminating different prognosis trajectories.

To shed light on this, we first examined *complete and replicable* regional early brain alterations in a large toddler sample (166 ASD, 109 TD). Specifically, we comprehensively and systematically investigated regional brain volume alterations and cortical surface area and thickness perturbations in ASD compared to TD toddlers. We then examined the replicability of these regional differences in an independent toddler cohort (38 ASD, 37 TD) using *the same preprocessing pipeline and the same statistical methods*. We further investigated whether these brain alterations were associated with contemporaneous behavioral manifestations of ASD quantified by symptom severity assessed using Autism Diagnostic Observation Schedule (ADOS), and cognitive and behavioral performance evaluated using Mullen and Vineland. Lastly, we investigated whether including brain size measures found to be altered at intake age would improve a model’s ability to predict language outcome at 3-4 years of age beyond intake clinical and behavioral measures.

## Materials and methods

This study was approved by the Institutional Review Board at the University of California, San Diego. Written informed consent was obtained from parents or legal guardians for all toddlers included in this study. Parents or legal guardians were compensated for their participation.

### Main sample

All toddlers were recruited through community referrals or a general population-based screening method called Get SET Early ^36^, also known as the 1-Year Well-Baby Check-Up Approach ^37,38^, allowing detection of ASD at early ages (e.g., ∼12 months). Toddlers were tracked from an intake assessment (1-3 years of age) and followed roughly every 12 months until 3 to 4 years of age (outcome visit). All toddlers participated in a series of clinical and behavioral assessments at each visit, including ADOS (Module T, 1, or 2) for ASD symptom evaluation ^39-41^, the Mullen Scales of Early Learning ^42^ for evaluating early cognition, and the Vineland Adaptive Behavior Scales ^43^ for assessing a child’s functional skills in four different developmental domains.

All assessments were performed by licensed psychologists with PhD degrees (e.g., C.C.B.) and occurred at UCSD Autism Center of Excellence. Diagnosis at the most recent clinical visit was used in this study. Diagnosis of ASD is determined by highly experienced and licensed psychologists (C.C.B.) using diagnostic criteria in DSM □ ^44^ or □ ^1^ in combination with the gold-standard ADOS evaluation ^45^. Typically developing (TD) toddlers showed no history of any developmental delay. Due to the fact that a large proportion of toddlers with ASD were scored at the floor of the standardized scores on Mullen subscales, we computed a ratio score for each subscale by dividing the age equivalent score by the toddler’s chronological age ^46-49^. We used these ratio scores to evaluate their associations with brain morphometry.

Clinical and behavioral scores and structural MRI (sMRI) scans were collected from 343 toddlers (198 ASD and 145 TD). Around 30% of toddlers had follow-up sMRI scans collected, contributing to 447 scans in total. Among 343 toddlers, 68 had poor sMRI scans or scans with bad segmentation quality (see details later) and were excluded from the study (75 scans were discarded), yielding data from 275 toddlers (166 ASD, 109 TD; 202 male, 73 female; 13–50 months old). Out of 275 toddlers, 187 had only an intake sMRI scan collected, 88 had one or more follow-up sMRI scans collected at intervals ranging from 0.5 to 27 (mean ± standard deviation:13.03 ± 3.35) months after the initial/previous scan, contributing to 372 scans in total. Demographic and clinical characteristics at the time of scanning of 275 toddlers are displayed in Table 1.

**Table 1.** Demographic Information and Clinical Test Scores for ASD and TD toddlers in Main Sample.

| Characteristics | ASD (166 toddlers) | TD (109 toddlers) | p value (ASD vs. TD) |
| --- | --- | --- | --- |
| **Demographics at MRI and clinical visit** |  |  |  |
| Sex (M/F) | 137/29 | 65/44 | $2.60 \times 10^{-5}$ <sup>a</sup> |
| Age at MRI scan (months) | 30.02 (8.31) | 24.63 (9.00) | $6.49 \times 10^{-7}$ <sup>b</sup> |
| Age at clinical visit (months) | 28.76 (8.25) | 23.09 (9.20) | $2.71 \times 10^{-7}$ <sup>b</sup> |
| **ADOS (module T, 1 or 2) score** |  |  |  |
| ADOS SA | 13.77 (4.43) | 1.96 (2.22) | $1.30 \times 10^{-69}$ <sup>c</sup> |
| ADOS RRB | 3.87 (1.93) | 0.28 (0.64) | $2.95 \times 10^{-46}$ <sup>c</sup> |
| ADOS Total | 17.64 (5.55) | 2.25 (2.45) | $6.04 \times 10^{-74}$ <sup>c</sup> |
| **Mullen score** |  |  |  |
| Ratio fine motor | 86.05 (17.40) | 111.93 (14.04) | $4.40 \times 10^{-17}$ <sup>c</sup> |
| Ratio visual reception | 87.25 (19.58) | 116.33 (16.82) | $7.59 \times 10^{-19}$ <sup>c</sup> |
| Ratio expressive language | 63.89 (22.32) | 104.46 (19.42) | $1.63 \times 10^{-30}$ <sup>c</sup> |
| Ratio receptive language | 64.43 (24.40) | 110.93 (19.84) | $1.73 \times 10^{-33}$ <sup>c</sup> |
| Early learning composite | 73.27 (17.80) | 111.75 (17.36) | $7.94 \times 10^{-35}$ <sup>c</sup> |
| **Vineland standard score** |  |  |  |
| Adaptive behavior composite | 80.44 (9.99) | 102.96 (10.07) | $4.73 \times 10^{-43}$ <sup>c</sup> |
| Daily living skills | 84.22 (11.20) | 103.26 (10.66) | $7.09 \times 10^{-29}$ <sup>c</sup> |
| Socialization | 81.05 (10.85) | 104.21 (9.64) | $2.36 \times 10^{-43}$ <sup>c</sup> |
| Motor skills | 90.61 (11.38) | 99.99 (8.97) | $2.12 \times 10^{-9}$ <sup>c</sup> |
| Communication | 77.45 (13.97) | 102.94 (11.08) | $5.20 \times 10^{-39}$ <sup>c</sup> |

<sup>a</sup>Pearson's chi-squared test.
<sup>b</sup>Welch's two sample t test.
<sup>c</sup>N-way ANOVA test including age and sex as covariates.
Note, values for age and all clinical test scores are presented as mean (SD). SD represents standard deviation. ADOS SA represents ADOS social affect, and ADOS RRB presents ADOS restricted and repetitive behavior.

### MRI data acquisition and preprocessing

Imaging data were collected on a 1.5T General Electric MRI scanner during natural sleep at night; no sedation was used. Structural MRI data were collected with a T1-weighted IR-FSPGR (inversion recovery fast-spoiled prepared gradient recalled) sagittal protocol with TE (echo time) = 2.8 ms, TR (repetition time) = 6.5 ms, flip angle = 12°, bandwidth = 31.25 kHz, field of view = 24 cm, and slice thickness = 1.2 mm. All sMRI scans were parcellated using FreeSurfer 5.3 (http://surfer.nmr.mgh.harvard.edu/) ^50^ based on the Desikan-Killiany atlas ^51^ to provide global and regional brain morphometric measures, including total brain volume, total surface area (SA), mean cortical thickness, cortical sub-regional volume/SA/thickness, and subcortical volumes.

FreeSurfer aligns each toddler’s brain to an average brain derived from cortical folding patterns through nonlinear surface-based registration ^52^. This tool has been validated for studies of children ^53^ and has shown great success in large pediatric studies ^35,54,55^. Quality evaluation was further performed on the raw and segmented sMRI scans by two independent raters (M.D. and K.C.) with a rating scale ranging from 0 to 3 (0=best, 1=great, 2=usable, 3=unusable). Out of 447 sMRI scans, 75 were rated as unusable and were excluded from the study, yielding 372 scans.

### Replication sample

76 toddlers (38 ASD and 38 TD) recruited in our previous study ^14^ were used as a replication sample. Toddlers were recruited through clinical referral or advertisements and were diagnosed by the same licensed psychologist (C.C.B) with the above-mentioned criteria. sMRI scans were collected at the same site with a 1.5T Siemens Symphony system during toddler’s natural sleep at night. A total of 170 sMRI scans were collected at intake and follow-up visits. All replication sMRI scans were preprocessed with FreeSurfer 5.3 using the *same pipeline* and *same Linux platform* as used for main samples. Similarly, sMRI scans with excessive motion or bad segmentation quality were excluded, yielding data from 75 unique toddlers (38 ASD, 37 TD; 55 male, 20 female) and 167 scans. The detailed participant recruitment, diagnosis evaluation, and scan collection can be found in ^14^.

### Brain structure difference between ASD and TD toddlers

For both the main and replication samples, ASD vs. TD differences in regional brain size was examined using the same linear mixed-effect models as described later. Brain global measures (the estimated total intracranial volume (eTIV), total cortical SA, and mean cortical thickness) differences between ASD and TD were tested using the model:

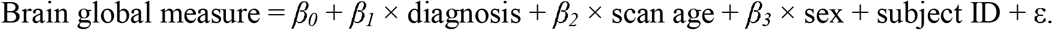

where each global brain measure was treated as the dependent variable, and fixed-effect predictors included diagnosis, age at scan, and sex. Subject was treated as a random effect to take longitudinal scans into account. Diagnosis was coded as a dummy variable (ASD = 1, TD = 0). Thus, for each brain region tested, the beta value of diagnosis can be interpreted as how much larger/smaller (unit: cm for thickness, cm^2^ for SA, cm^3^ for volume) ASD toddlers’ brains are compared to TDs’ brains. ASD vs. TD differences in cortical and subcortical volume, cortical regional surface area and thickness were tested using the linear mixed effect model as below:

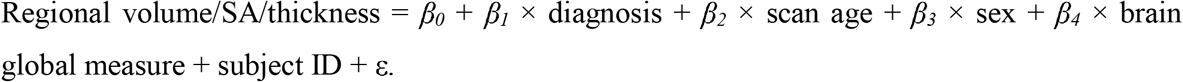

where volume/SA/thickness of each brain region was treated as the dependent variable. Subject was treated as a random effect, and other predictors (diagnosis, age at scan, sex, and brain global measure) were modeled as fixed effects. Brain global measures included eTIV for testing sub-cortical and cortical regional volume, total cortical SA for testing regional SA, and mean cortical thickness for testing regional thickness measures. To identify cortical regions with significant volume/SA/thickness differences between ASD and TD in the main sample, a false discovery rate (FDR) at *p* < 0.05 was applied to correct for 68 comparisons (68 cortical regions in left (LH) and right (RH) hemispheres). FDR at *p* < 0.05 was also applied to correct for comparisons of subcortical regions, cerebellum (LH and RH), and corpus callosum (CC) regions separately. The identified ASD vs. TD differences were considered as replicated if the corresponding *p* values were less than 0.05 in the replication sample.

### Brain-behavior association analyses

Associations between behavioral measures (ADOS, Mullen, and Vineland) evaluated at the time of scan and brain regions showing significant ASD vs. TD differences were examined in ASD and TD toddlers separately using the linear regression model:

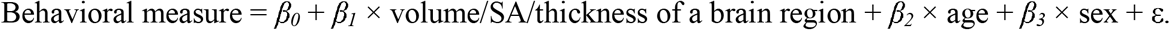

where each behavioral measure is treated as the response variable, and age, sex, and volume/SA/thickness of a brain region were predictors. FDR at *p* < 0.05 was applied to correct for multiple comparisons. Moreover, brain-by-diagnosis interaction effects in predicting behavioral measures were investigated using the regression model:

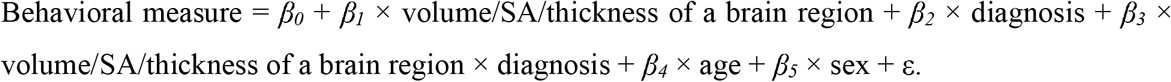

### Predicting language outcome for ASD toddlers

Language outcome of ASD toddlers was stratified as ASD Good or ASD Poor based on Mullen EL and RL T scores at outcome visit, as previously employed for prognostic analyses ^33-35^. An ASD toddler was grouped as ASD Poor if both Mullen EL and RL T scores were below -1 SD of the T score norm of 50 (i.e., T < 40). An ASD toddler was classified as ASD Good if the toddler had either Mullen EL or RL T scores equal to or greater than -1 SD of the normative T score of 50 (i.e., T ≥ 40). Out of 166 ASD toddlers, 157 had a Mullen evaluation at outcome visit and were stratified into two outcome groups: ASD Good (N = 69; 59 males, 10 females; age = 33.88 ± 4.44 months) and ASD Poor (N = 88; 71 males, 17 females; age = 34.55 ± 5.18 months). These 157 ASD toddlers were further used for language outcome prediction analysis.

To predict language outcome (ASD Good/Poor), we employed the support vector machine (SVM) with ridge regularization. SVM with ridge can select features of importance to achieve a stable classification result. We tested and evaluated three different models: clinical/demographic only, sMRI only, and clinical/demographic + sMRI models. The clinical/demographic only model used behavioral (ADOS, Mullen and Vineland) and demographic (sex, age at intake, and gap between intake and outcome visit) variables at intake visit. The sMRI only model leveraged age and sex-adjusted intake FreeSurfer measures (age and sex effects were estimated using TD data ^56^) within regions that showed significant ASD vs. TD differences. The clinical/demographic + sMRI model used all intake features included in clinical/demographic only and sMRI only models. Each variable/feature was scaled to be between 0 and 1 prior to SVM for all models. Each model was cross-validated with the training samples using 5-fold cross-validation, and its performance was evaluated with a hold-out testing set. Among 157 ASD toddlers, 125 (80% samples, ASD Good/Poor = 56/69; female/male = 21/104, age: 34.22 ± 4.81 months) were utilized for training, with the remaining 32 participants (20% samples, ASD Good/Poor = 13/19; female/male = 6/26, age: 34.40 ± 5.13 months) used as a hold-out testing set. Accuracy, sensitivity, specificity, and area under the receiver operating characteristic (ROC) curve (AUC) were computed to reflect the performances of prediction models.

### Data availability

Raw sMRI and clinical data are available from the National Institute of Mental Health Data Archive (NDA, collection ID = 9). The processed data and code are available from the authors upon reasonable request.

## Results

### ASD vs. TD brain structure difference in main sample

In the main sample, no significant ASD vs. TD difference was observed for eTIV (*p* = 0.96), total cortical volume (*p* = 0.07), total cortical SA (*p* = 0.49), or mean cortical thickness (*p* = 0.47). However, ASD poor toddlers presented significantly greater total cortical volume compared to TD (*p* = 2.56×10^−3^, Cohen’s d (referred as d hereafter) = 0.39, beta = 8.62). Four cortical regions showed significant volume differences between ASD and TD toddlers after FDR at *p* < 0.05 correction (Fig. 1 upper left): ASD toddlers had significantly *increased* gray matter volume (GMV) in LH fusiform (*p* = 2.44×10^−4^, d = 0.42, beta = 0.49), LH (*p* = 9.23×10^−4^, d = 0.37, beta = 0.47) and RH (*p* = 6.22×10^−5^, d = 0.45, beta = 0.59) middle temporal regions compared to TD toddlers; ASD toddlers also showed significant regional GMV *reduction* in RH caudal anterior cingulate compared to TD (*p* = 6.12×10^−4^, d = -0.39, beta = -0.19). Moreover, six cortical regions showed a significant thickness difference between ASD vs. TD toddlers (Fig. 1 upper right). Compared to TD, ASD toddlers had significantly *thicker* cortex in LH superior temporal (*p* = 9.22×10^−5^, d = 0.44, beta = 5.30×10^−3^) and RH banks of the superior temporal sulcus (bank STS) (*p* = 1.50×10^−3^, d = 0.36, beta = 7.08×10^−3^) regions, and significantly *thinner* cortical in LH caudal middle frontal (*p* = 2.76×10^−3^, d = -0.34, beta = -3.94×10^−3^), LH (*p* = 1.76×10^−3^, d = -0.35, beta = -4.48×10^−3^) and RH pars opercularis (*p* = 8.18×10^−4^, d = -0.38, beta = -5.39×10^−3^), as well as LH pericalcarine (*p* = 4.30×10^−3^, d = -0.32, beta = -6.12×10^−3^) regions. ASD toddlers showed significantly *reduced* cortical SA compared to TD in RH caudal anterior cingulate (*p* = 3.04×10^−5^, d = -0.47, beta = -0.46), RH medial orbitofrontal (*p* = 5.28×10^−5^, d = -0.46, beta = -0.57) and RH posterior cingulate (*p* = 3.45×10^−4^, d = -0.41, beta = -0.43) regions (Fig. 1 lower left). Outside of the cortex, ASD toddlers also presented significantly *increased* volume compared to TD in posterior CC (*p* = 4.05×10^−3^, d = 0.33, beta = 0.04), mid posterior CC (*p* = 0.03, d = 0.24, beta = 0.01), and mid anterior CC (*p* = 0.02, d = 0.26, beta = 0.02), and *decreased* GMV in right cerebellum GMV (*p* = 6.74×10^−3^, d = -0.31, beta = -1.18) (Fig. 1 lower right). No significant ASD vs. TD difference was observed for subcortical regional GMV (*p* > 0.05 for all subcortical regions). Violin plots of brain regions showing significant ASD vs. TD difference are displayed in supplemental Fig. S1. Results of brain regions showing nominal significant ASD vs. TD difference (*p* < 0.05) are listed in the supplemental spreadsheet.

**Figure 1.**
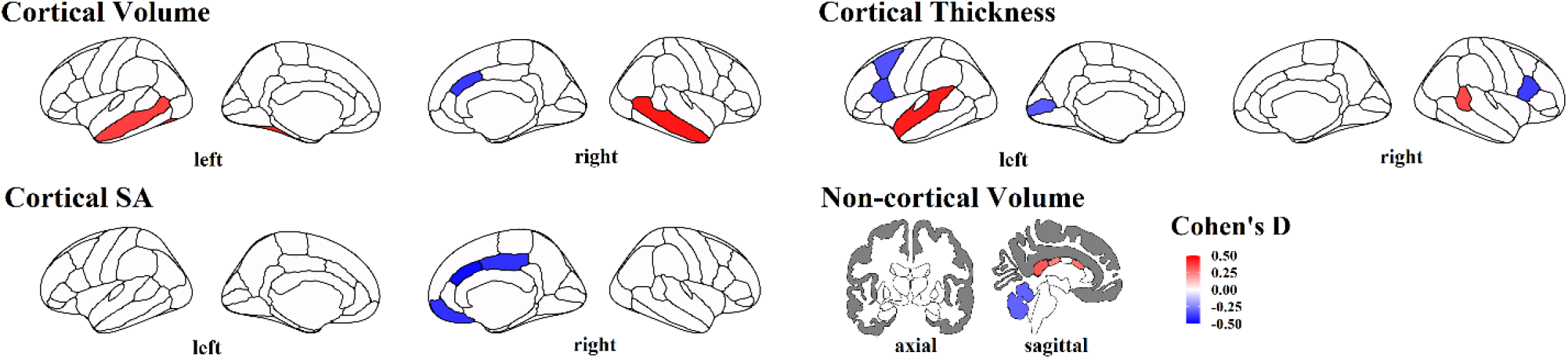
Brain regions showing significant differences between ASD and TD toddlers in the *main* sample in terms of cortical volume, non-cortical volume, cortical thickness and cortical SA. Colors represent corresponding effect sizes (Cohen’s D), where regions with hot colors showed significant *increases* in size among ASD compared to TD and regions with cold colors showed significant *decreases* in size among ASD compared to TD; the darker the color, the larger the difference between ASD and TD.

### ASD vs. TD brain structure difference in replication sample

In the replication sample, ASD toddlers had significantly bigger brains (*p* = 0.04, d = 0.41, beta = 1.15), greater total cortical volume (*p* = 1.72 × 10^−3^, d = 0.60, beta = 17.22) and larger mean cortical thickness (*p* = 1.42 × 10^−4^, d = 0.72, beta = 7.87 × 10^−3^) compared to TD. No significant ASD vs. TD difference was observed for total cortical SA (*p* = 0.13). Three out of four cortical regions showing significant GMV differences between ASD and TD toddlers in the main sample were replicated (Fig. 2 left): ASD toddlers had significantly *increased* GMV in LH fusiform (*p* = 0.03, d = 0.43, beta = 0.46), LH (*p* = 5.78×10^−8^, d = 0.95, beta = 1.22) and RH (*p* = 1.71×10^−4^, d = 0.64, beta = 0.94) middle temporal regions compared to TD toddlers. Among 6 regions showing significant thickness differences between ASD and TD toddlers in the main sample, three were replicated (Fig. 2 middle): Compared to TD, ASD toddlers had significantly *thicke* cortex in LH superior temporal (*p* = 1.32×10^−6^, d = 0.85, beta = 1.04×10^−2^) and RH bank STS (*p* = 0.05, d = 0.33, beta = 6.56×10^−3^) regions, and significantly thinner cortex in LH pars opercularis (*p* = 1.49×10^−3^, d = -0.54, beta = -6.89×10^−3^) region. Moreover, ASD toddlers showed significantly *increased* volume of mid anterior CC (*p* = 2.85×10^−3^, d = 0.50, beta = 0.03), but *decreased* volume in right cerebellum cortex (*p* = 1.56×10^−2^, d = -0.41, beta = -1.82) compared to TD (Fig. 2 right). None of the three cortical regions showing significant SA differences were replicated (*p* > 0.05). Violin plots of brain regions that were replicated for ASD vs. TD differences are presented in supplemental Fig. S2.

**Figure 2.**
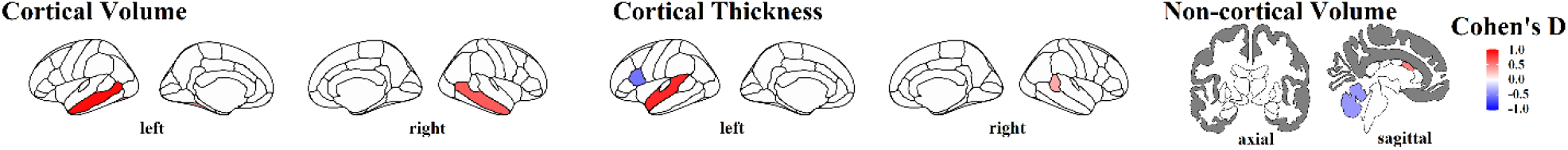
Brain regions replicated for ASD vs. TD differences in cortical volume, non-cortical volume and cortical thickness. Colors represent corresponding effect sizes (Cohen’s D), where regions with hot colors showed significant *increases* in size among ASD compared to TD and regions with cold colors showed significant *decreases* in size among ASD compared to TD; the darker the color, the larger the difference between ASD and TD.

### Associations between brain size and behavior

Among 13 regions showing significant ASD vs. TD differences in the main sample, four were significantly related to ADOS symptom severity or Mullen subscale scores after FDR correction (Fig. 3 and Fig. S3). In ASD toddlers (Fig. 3), *larger* (i.e., more aberrant) GMV in LH fusiform was significantly associated with *higher* ADOS total (i.e., more severe symptoms, *r* = 0.17, *p* = 2.62 × 10^−2^), and *lower* Mullen ELC (*r* = -0.25, *p* = 1.51 × 10^−3^), *lower* Mullen ratio RL (*r* = -0.28, *p* = 2.76 × 10^−4^) and *lower* Mullen ratio VR scores (*r* = -0.26, *p* = 9.58 × 10^−4^) (i.e., poorer performance on Mullen subscales). Similarly, *larger* (i.e., more aberrant) GMV in LH and RH middle temporal was significantly associated with *higher* ADOS SA (LH: *r* = 0.24, *p* = 1.74 × 10^−3^; RH: *r* = 0.24, *p* = 2.30 × 10^−3^) and *higher* ADOS total (LH: *r* = 0.24, *p* = 1.79 × 10^−3^; RH: *r* = 0.24, *p* = 2.06 × 10^−3^) scores, and *lower* Mullen ELC (LH: *r* = -0.19, *p* = 1.87 × 10^−2^; RH: *r* = -0.20, *p* = 9.94 × 10^−3^), *lower* Mullen ratio RL (LH: *r* = -0.19, *p* = 1.94 × 10^−2^; RH: *r* = -0.20, *p* = 1.15 × 10^−2^) and *lower* Mullen ratio VR (LH: *r* = -0.19, *p* = 1.86 × 10^−2^; RH: *r* = -0.18, *p* = 2.35 × 10^−2^). Paradoxically, *larger* (i.e., less aberrant) SA in RH caudal anterior cingulate was significantly related to lower Mullen ratio RL (*r* = -0.22, *p* = 6.05 × 10^−3^). In TD toddlers (Fig. S3), only *larger* GMV in LH middle temporal was significantly associated with *higher* Mullen ratio VR scores (*r* = 0.23, *p* = 0.02), opposite of the direction of association seen in ASD.

**Figure 3.**
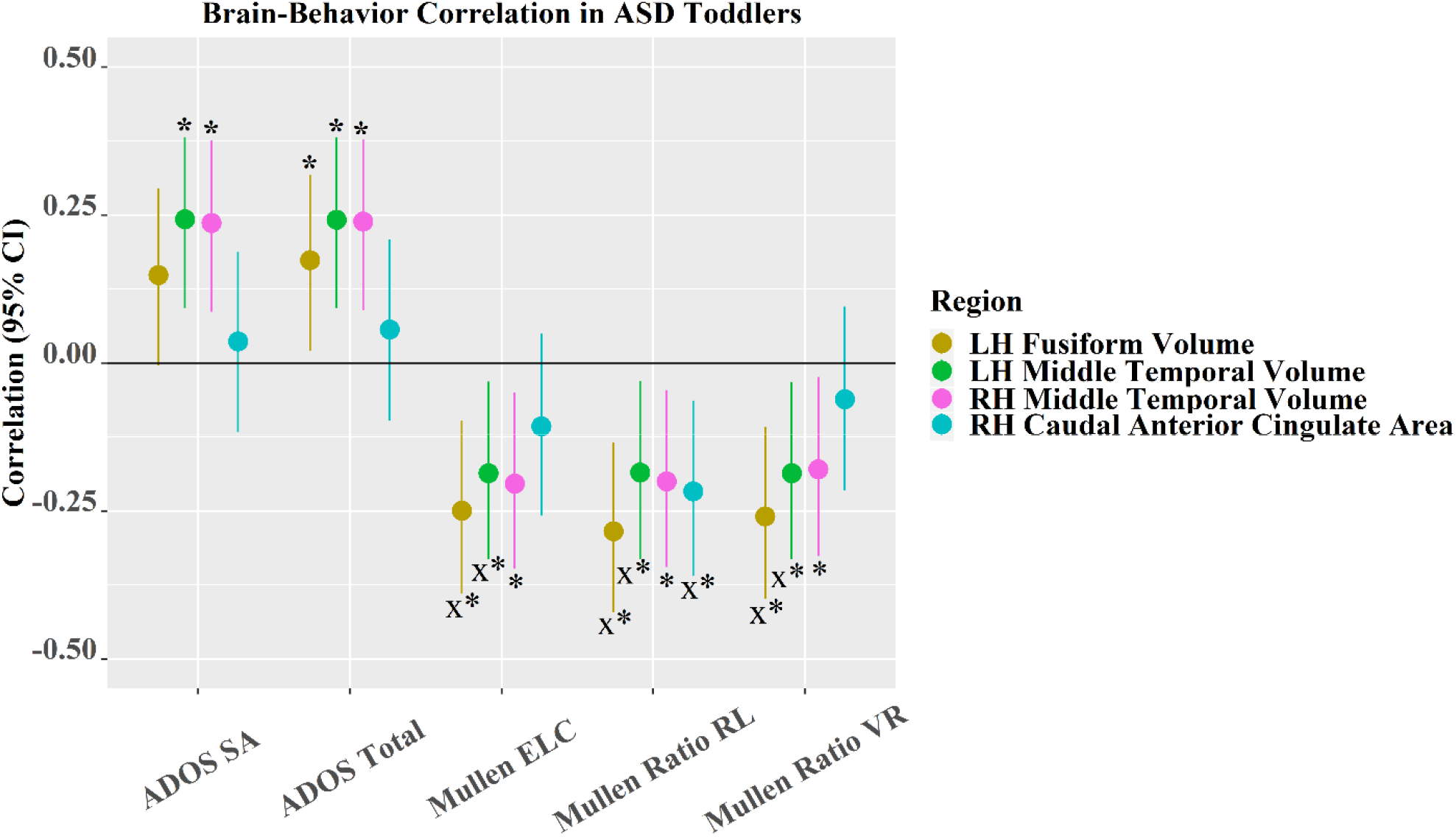
Brain-behavior correlation and its 95% CI in ASD toddlers. Note that * indicates the correlation is significant; x represents that brain measures significantly interact with diagnosis (ASD/TD) to predict behavior; colors of medium dark shades of yellow, green, cyan and a medium light shade of magenta denote LH fusiform volume, LH middle temporal volume, RH caudal anterior cingulate SA, and RH middle temporal volume, respectively.

Importantly, GMV in LH fusiform and LH middle temporal (MT) significantly interacted with diagnosis to predict Mullen ELC (fusiform: *p* = 0.01; MT: *p* = 2.73 × 10^−3^), Mullen ratio RL (fusiform: *p* = 3.74 × 10^−3^; MT: *p* = 1.18 × 10^−2^), and Mullen ratio VR (fusiform: *p* = 2.33 × 10^−3^; MT: *p* = 2.79 × 10^−4^). Associations were strongly negative in the ASD group, but near zero or positive in the TD group. Moreover, SA in RH caudal anterior cingulate significantly interacted with diagnosis to predict Mullen ratio RL (*p* = 1.52 × 10^−2^). Scatter plots of significant brain-behavior associations are presented in Fig. S4.

### Predicting language outcome for ASD toddlers

Mullen ELT and RLT scores of TD and ASD toddlers with good/poor language outcome are displayed in Fig. S5, where ASD Good toddlers showed similar language outcome as TD toddlers. Fig. 4 plots the performance of clinical/demographic only, sMRI only, and clinical/demographic + sMRI models for classifying ASD Good versus Poor language outcome. Sensitivity and specificity reflect the accuracy for correctly detecting ASD Poor and ASD Good, respectively. Combining intake clinical/demographic and sMRI features yielded the highest accuracy (81%) and AUC (79%) compared to that from a single modality (sMRI only model: accuracy = 69%, AUC = 63%; clinical/demographic only model: accuracy = 72%, AUC = 70%). The clinical/demographic only model achieved slightly higher accuracy than the sMRI only model, especially for detecting ASD Good toddlers (i.e., specificity). sMRI had the highest accuracy in detecting ASD Poor toddlers (i.e., sensitivity). Fig. S6 displays the contribution (weight) of each intake clinical/demographic and sMRI feature to predicting the language outcome of ASD toddlers.

**Figure 4.**
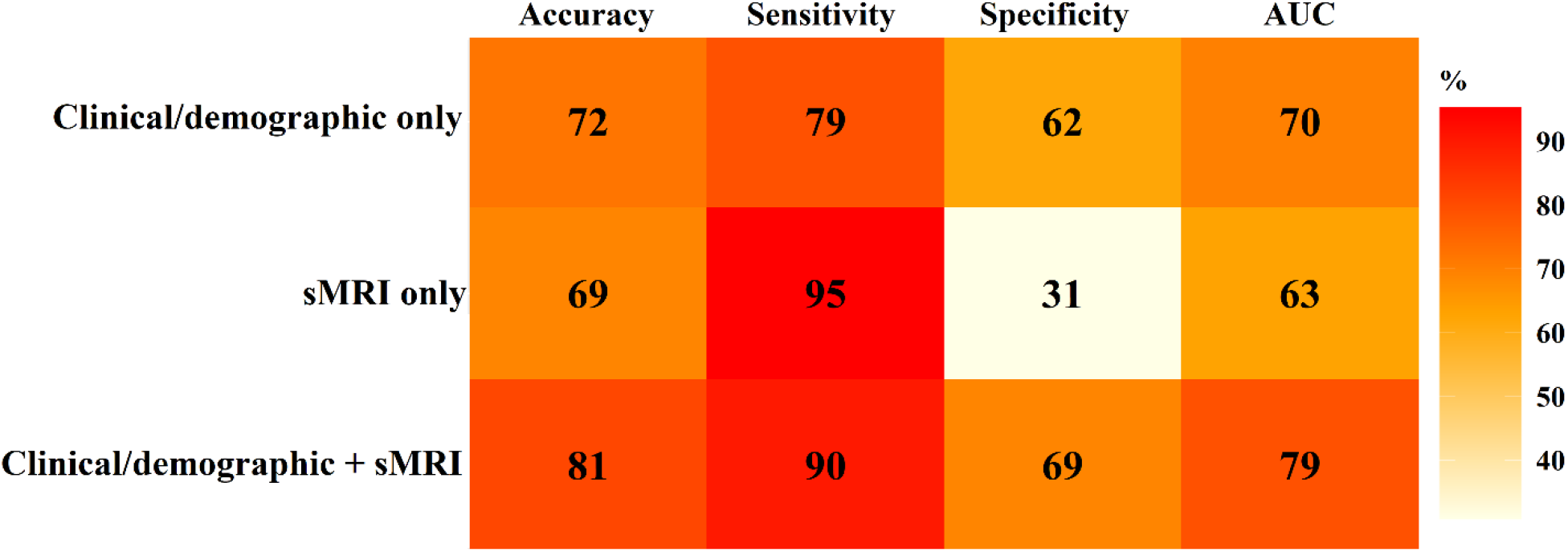
Accuracy, sensitivity (for detecting ASD Poor), specificity (for detecting ASD Good), and AUC values of clinical/demographic only, sMRI only, clinical/demographic + sMRI models for predicting ASD Good versus Poor language outcome. Features used in each model were collected at the intake visit (the earliest clinical visit, mean age = 2.5 years). Language outcome was evaluated at 3-4 years of age.

## Discussion

In this study, we surveyed the volume, thickness, and surface area of all regions across the brain to observe which size measures were reproducibly altered in ASD toddlers compared to TD toddlers. Identified brain regions are mainly involved in receptive and expressive language, social and face processing (bank STS, middle temporal, superior temporal, medial orbitofrontal, caudal anterior cingulate, posterior cingulate, pars opercularis, caudal middle frontal)^57-67^. Additional regions included those involved in motor, behavioral, cognitive, and language control; primary visual processing and interhemispheric communication (cerebellum; primary visual cortex, corpus callosum)^68-75^. Morphometrically, we observed alterations in regional volume, thickness, and surface area relative to global measures. Thus, by first factoring out brain size, differentially increased or decreased growth in different anatomic measures in ASD-relevant language, social, face processing and behavior regulation regions were differentially isolated and highlighted. Cortically, frontal lobe and midline structures tended to be smaller or thinner in ASD than TD; lateral temporal regions tended to be larger or thicker in ASD. Outside the cortex, larger callosal subregion volume and smaller cerebellum were observed. The majority of the identified GMV and cortical thickness alterations were replicated in an independent cohort. Importantly, larger (i.e., more aberrant) GMV in LH fusiform, LH and RH middle temporal were related to more severe ADOS symptoms and/or poorer Mullen cognitive (ELC, ratio RL, and ratio VR) performance in ASD toddlers. These relationships were significantly stronger in the ASD compared to TD group. Of clinical relevance, the identified brain features measured at intake (mean age = 2.5 years), when included in a predictive model along with clinical and demographic features, markedly improved the accuracy for classifying good vs. poor language outcome for toddlers with ASD at 3-4 years of age.

The identified regional alterations were largely consistent with previous findings. Studies have found that young children ^2,14,15,17^, adolescents, and adults ^76^ with ASD show GMV enlargement in the temporal lobe, especially in the superior, middle temporal and fusiform gyri ^77^. Increased cortical thickness in left hemisphere superior temporal cortex (LH STC) also appears to be a very strong and replicable finding in the literature, as evident in other large-scale studies in primarily adolescents and adults ^78,79^. The current results showcase that increased LH STC thickness is present even earlier in ASD in toddlerhood and with larger effect sizes than studies in older ASD individuals. This developmentally-ubiquitous increase in cortical thickness of LH STC may yield insight about early development processes that contribute to cortical thickness (e.g., proliferation of excitatory neuronal cell types in different cortical layers). Furthermore, given that normative brain charts indicate that cortical thickness tends to peak in early childhood followed by slow decline over the lifespan ^80^, and so, these ASD toddler results combined with others in older ASD samples would indicate that increased early developmental cortical thickening combined with attenuated cortical thinning of LH STC may be a robust and key neural feature of ASD neurodevelopment. Given the observations of early developmental functional abnormalities in LH STC for language ^33,34,81^, these converging results may implicate that atypical structural development and underlying genomic mechanisms affecting LH STC ^34,35^ may perturb the ability of this region to develop functional specialization for processes like language and social-communication.

GMV reduction in the cerebellum has been well-documented for individuals with ASD spanning from childhood to late adulthood ^25-27^. Postmortem studies also revealed that subjects with ASD showed decreased number ^82^ and reduced size ^83^ of Purkinje cells in the cerebellar hemisphere and vermis. The identified volume increase in CC in ASD toddlers aligned with the finding that infants with ASD had significantly increased SA and thickness in CC starting at 6 months of age, and the increase was particularly robust in the anterior CC at both 6 and 12 months ^22^. Other studies ^21-24^ suggest that CC in individuals with ASD likely undergoes overgrowth at early ages ^22^, followed by abnormally slow or arrested growth, and later shows a reduction in adulthood ^23,24^. Our results of SA reduction in the orbitofrontal cortex and posterior cingulate were supported by a recent study led by Ecker ^84^. Moreover, the identified alterations in thickness aligned with the finding by Zielinski et. al. ^85^ that individuals with ASD showed reduced thickness in the bilateral caudal middle frontal and the left pars opercularis during childhood and adolescence as well as in the right pars opercularis during adulthood.

By first factoring out brain size, we revealed abnormal cortical patterning in multiple ASD-relevant language, social, face processing and behavior regulation regions. This abnormality was manifest in a complex map of *differentially* increased or decreased GM volume, surface area and thickness and highlights the presence of dysregulated cortical growth. These different early-age regional alterations of cortical attributes may be the result of progressive dysregulation in multiple neural processes and stages, consistent with prenatal multi-process, multi-stage models of ASD ^86,87^. This advances our recent finding of atypical anterior-posterior and dorsal-ventral genetical cortical patterning in ASD toddlers with poor language and social outcomes^35^. In that study, atypical gene co-expression included genes involved in prenatal cortical patterning; all progenitor cell types involved in symmetrical and asymmetrical cell division that can alter surface area and cortical thickness; and excitatory neurons, oligodendrocyte precursors, endothelial cells, and microglia that may affect thickness. Thus, effects span multiple prenatal stages and growth processes, that we hypothesize lead to the multiple growth deviances in volume, surface area, and thickness across key cortical regions that we report here.

One mechanism that could be involved in these effects is the overactivity of a prenatal multi-pathway gene network, a gene dysregulation presented in ASD-derived prenatal progenitors and neurons and related to ASD social symptom severity^88^. This gene network, the DE-ASD Network, is composed of differentially expressed (DE) genes in ASD toddlers, and includes PI3K-AKT, RAS-ERK, Wnt, and Insulin receptor signaling pathways and upstream regulatory ASD risk genes. These signaling pathways normatively have a strong impact on prenatal brain patterning and development because they regulate proliferation, neurogenesis, differentiation, migration, neurite outgrowth, and synaptogenesis^87,89-93^. The Overactivity of gene expression in this DE-ASD Network is present in ASD vs. typical toddler progenitors and neurons and is greater in ASD toddlers who have more severe social symptoms^88^. Based on BrainSpan data (http://www.brainspan.org), this network normatively expresses during the first and second trimesters in multiple cortical areas during cortical patterning and progenitor cell division and neurogenesis^88^. Future studies should focus on the relationships between gene dysregulation in this DE-ASD Network in living ASD toddlers, brain cortical organoid models, and the toddlers’ neural and clinical phenotype to test this potential mechanism.

We found that toddlers with ASD who had more aberrant brain measures also showed more severe symptoms and poorer cognitive performance. The identified brain-behavior associations largely aligned with previous findings. Recently, Grecucci and colleagues ^31^ reported that larger GMV in an autism-specific structural network (including fusiform and middle temporal gyri) was related to higher ADOS subscales (social affect and restricted and repetitive behavior) and total scores. Rojas et. al. ^25^ also reported that GMV in the temporal region was positively associated with ADI-R Social and Communication total score. A study led by Dziobek reported that increased cortical thickness in the fusiform gyrus was related to impairments in face processing in individuals with ASD ^32^, consistent with our result that fusiform GMV was negatively related to the Mullen ratio VR score.

The identified brain regions were highly valuable for characterizing prognosis. The sMRI-clinical/demographic combined model achieved the highest accuracy for classifying ASD Good vs. ASD Poor, which was consistent with our previous finding that a multimodal model outperformed any single modality model ^33^. Integrating multiple modalities can take full advantage of both modality-unique and complementary information from other modalities that is key for parsing ASD heterogeneity. Notably, though sMRI model had the highest accuracy (sensitivity) for detecting ASD Poor, the accuracy for ASD Good was low. There were two possible reasons: 1) our samples included more ASD Poor than ASD Good toddlers, resulting in better detection of ASD Poor characteristics than that of ASD Good; and 2) the features input to SVM were more pronounced in ASD Poor than ASD Good in general (See supplemental Tables S1-S3), although a few showed reversed patterns.

The findings presented in this study should be considered in context with its strengths and limitations. Using brain regions showing significant ASD vs. TD differences as input for SVM reduced the likelihood of overfitting of the model. However, we may have missed other features that were important for discriminating ASD Good from ASD Poor. Future research should include a full exploration of all FreeSurfer features and training a more comprehensive model to improve the accuracy for detecting ASD Good. Another limitation is that while a majority of the identified brain alterations were replicated, further replication with larger samples is still necessary, especially for regions showing SA differences.

In summary, ASD toddlers showed GM alterations in regions mainly involved in language, social, face processing, and primary visual cortex. Most of the identified GM alterations were replicated in an independent cohort. Moreover, the identified GM alterations were associated with greater ASD symptom severity and cognitive impairments and showed great potential as prognostic biomarkers for language outcome prediction.

## Data Availability

Raw structural MRI and clinical data used in this study are available from the National Institute of Mental Health Data Archive (NDA, collection ID = 9). The processed data and code are available from the authors upon reasonable request.

https://nda.nih.gov/edit_collection.html?id=9

## Acknowledgments

We would like to thank the parents and children in San Diego who participated in our research, without whom this would not be possible. We are also fortunate to work with wonderful pediatricians and family practice physicians spanning a range of medical groups, including UCSD, Sharp Rees-Stealy, Scripps, Rady-Children’s Primary Care Medical Group, Chula Vista Pediatrics, Graybill Medical Group, Grossmont Pediatrics, Linda Vista Health Care Center, Mills Pediatrics, North County Health Services, San Diego Family Care, and Sea Breeze Pediatrics. We are grateful for their support.

## Author contributions

K.D. and E.C. conceptualized the study. K.D. performed data analysis and wrote the manuscript. E.C., L.E., M. V. L., and K. P. helped interpret the results and revise the manuscript. D. H. helped with sMRI data segmentation. M. D. and K.C helped with quality control on raw and segmented sMRI data. E.C., L.E., K.C., C.C.B, S.A., S. N. contributed to data collection and data management. E.C., K. P., L.E. and M. V. L. contributed to funding acquisition. All authors contributed to interpreting the results and discussion.

## Supplementary material

