## Supplemental Material for "Language and Social Regions Are Affected in Toddlers with Autism and Predict Later Language Outcome"

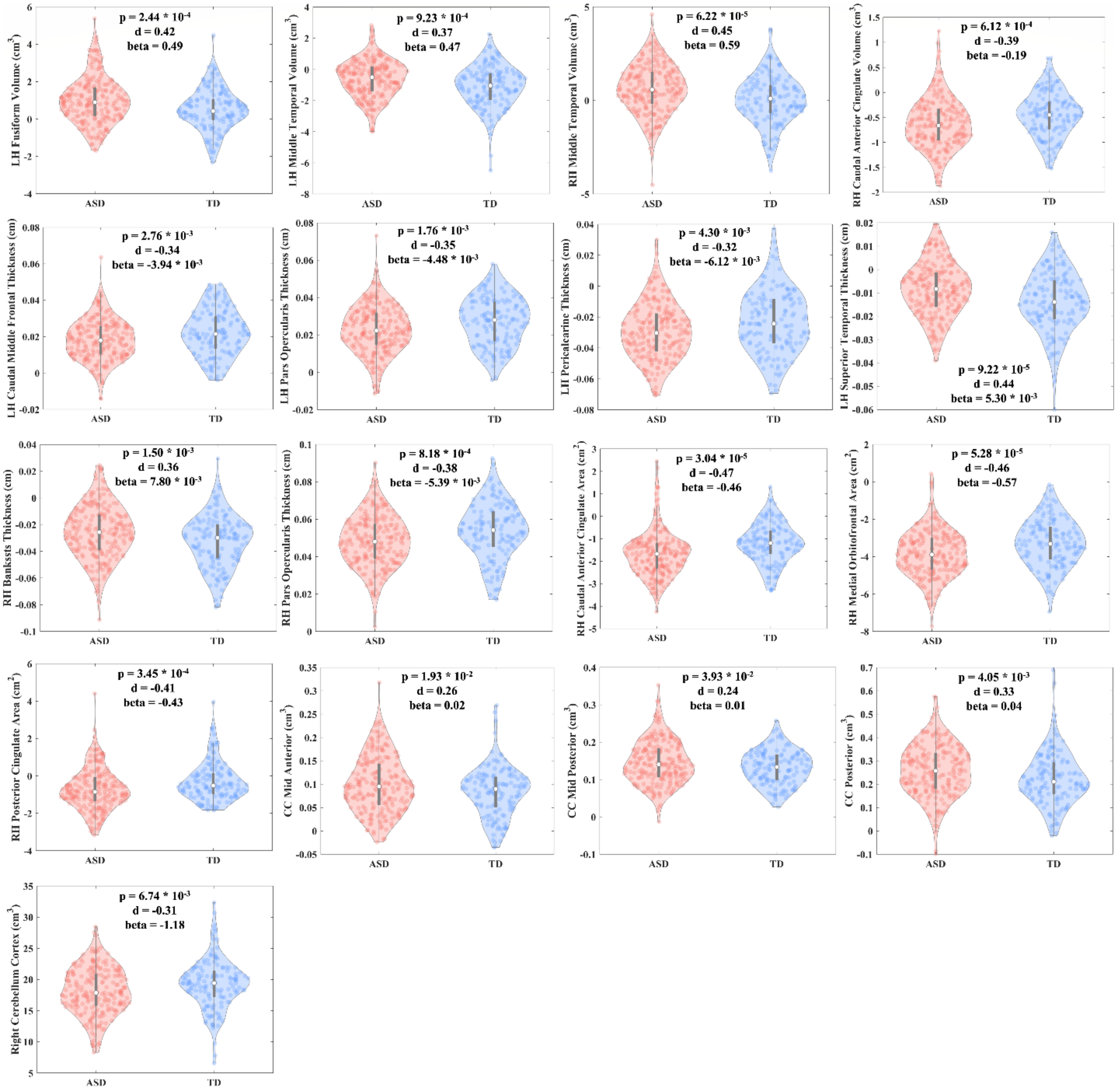


**Figure S1 Violin plots of brain regions showing significant difference between ASD and TD toddlers in *discovery* samples.** P, Cohen's d and beta values of ASD vs TD difference are displayed for each brain region. ASD and TD toddlers were presented as medium light shades of red and cyan-blue, respectively (the same for Figures S2, S4, and S5). Note that the brain measure of a specific region displayed on y axis was adjusted for the fixed effect from age and sex and the random effect from longitudinal scans (the same for Figure S2). Given that we coded diagnosis as a dummy variable (ASD = 1, TD = 0), the beta value for diagnosis can be interpret as how much larger/smaller (unit: cm for thickness, $cm^2$ for SA, $cm^3$ for volume) ASD toddlers' brain is compared to TDs' in a specific region (the same for Figure S2).

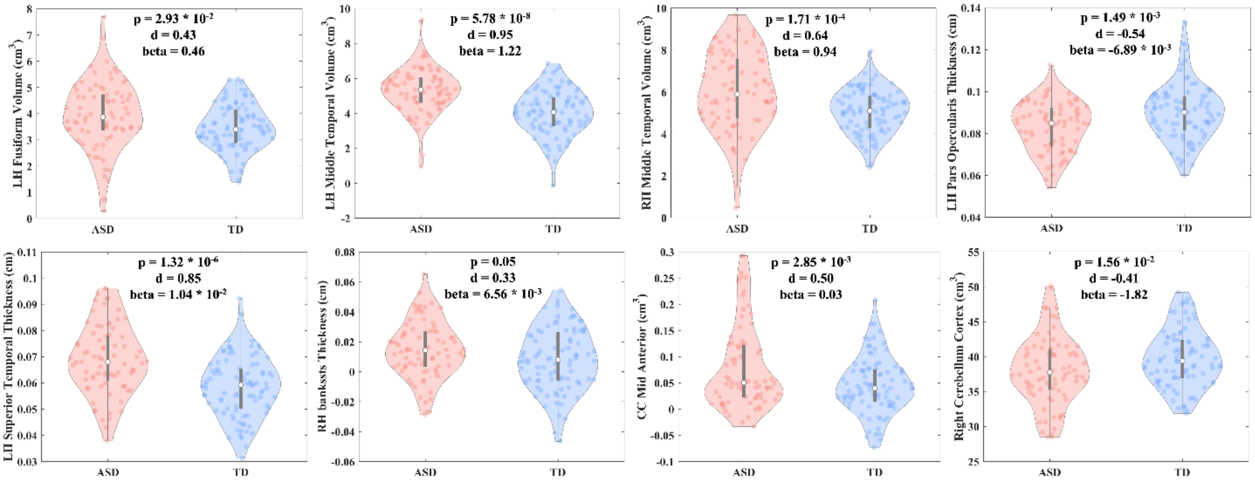


**Figure S2 Violin plots of brain regions that were replicated for ASD vs TD differences in *replication* samples.** P, Cohen's d and beta values of ASD vs TD difference are also presented for each brain region.

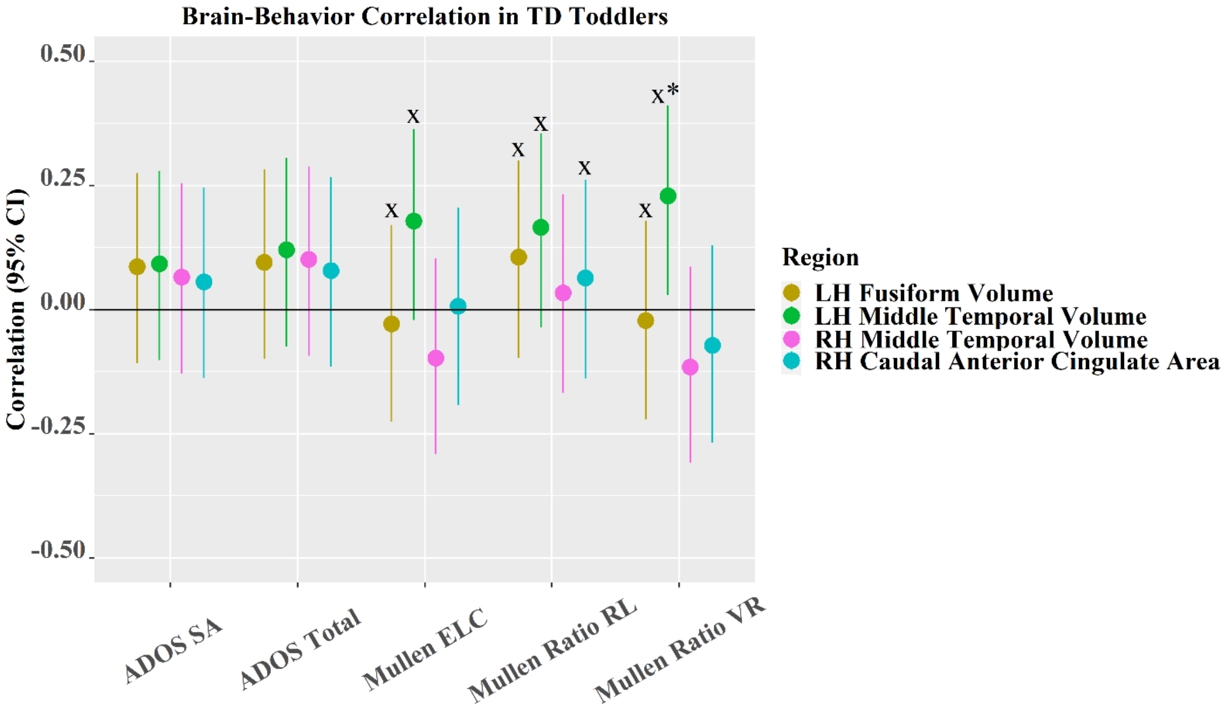


**Figure S3 Brain-behavior association and its 95% CI in TD toddlers.** Note that * indicates the correlation is significant; x represents that brain measures significantly interact with diagnosis (ASD/TD) to predict behavior; colors of medium dark shades of yellow, green, cyan and a medium light shade of magenta denote LH fusiform volume, LH middle temporal volume, RH caudal anterior cingulate SA, and RH middle temporal volume, respectively.

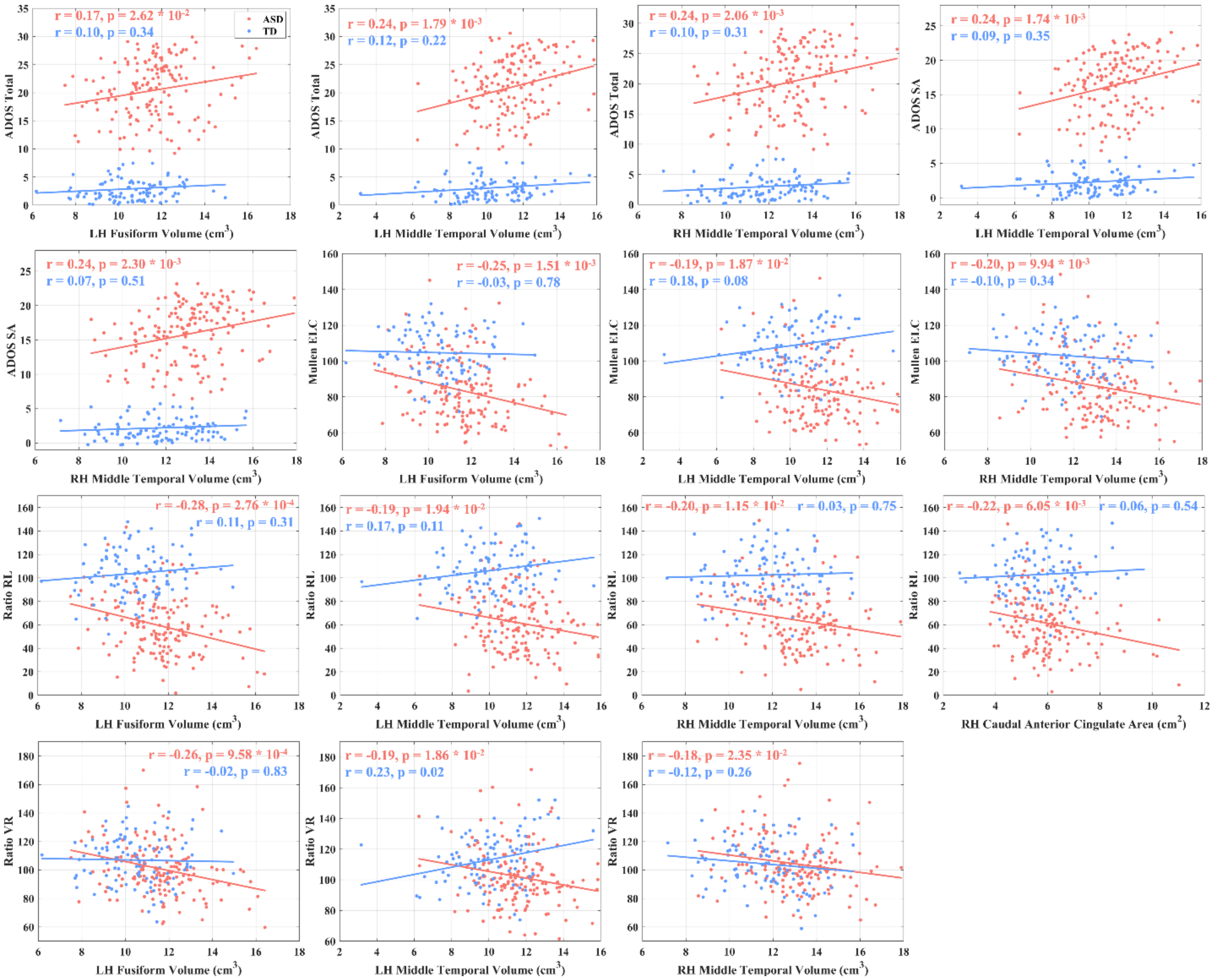


**Figure S4 Scatter plots for significant brain-behavior associations.** X axis represents a brain measure (volume/SA in a specific brain region) and y axis represents a behavioral measure (e.g., ADOS total, ADOS SA, Mullen ELC, Mullen ratio RL, and Mullen ratio VR). Note that the behavioral measure displayed on y axis was adjusted for age and sex effects.

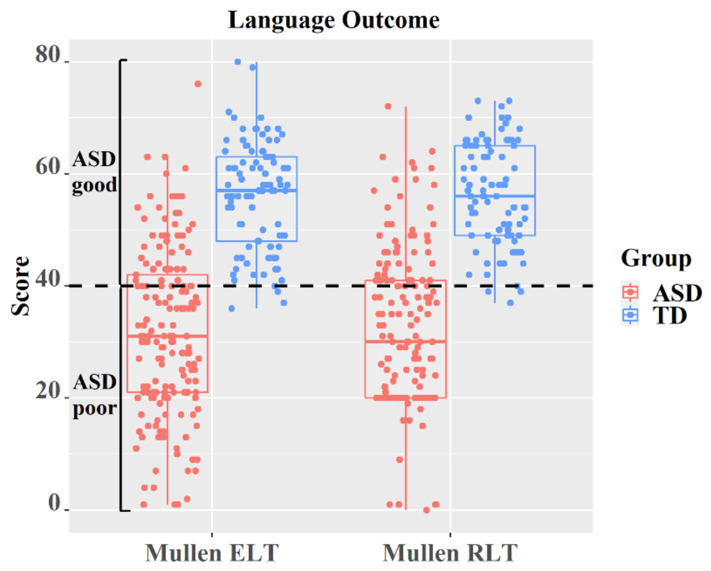


**Figure S5 Box plot of Mullen ELT and RLT scores of TD and ASD toddlers with good/poor language outcome.** The dash line indicates a score of 40 for Mullen ELT and RLT. Importantly, ASD good toddlers had similar language outcome as TD toddlers.

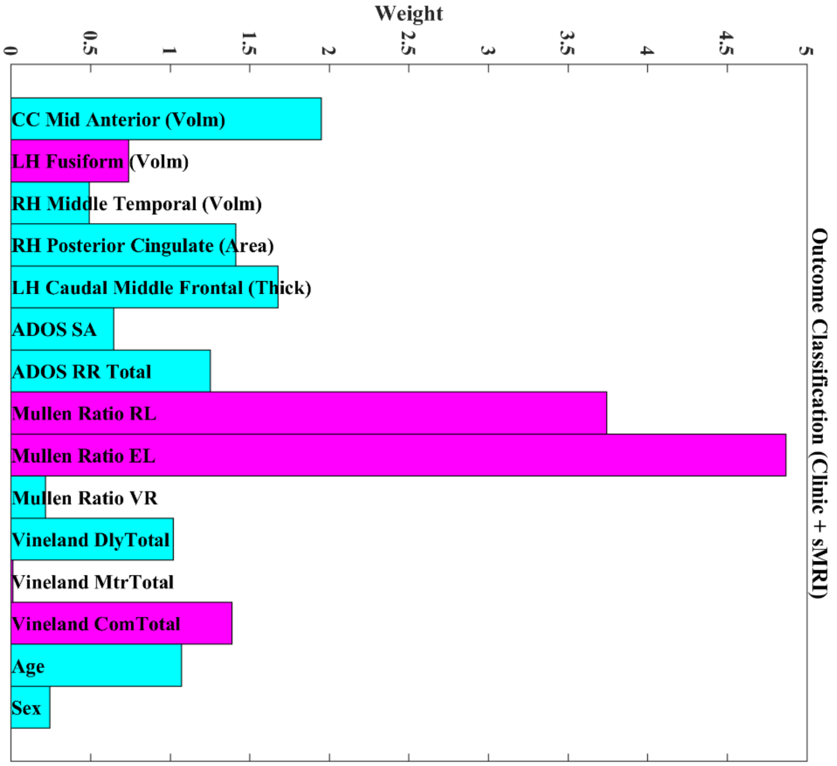


**Figure S6 Weights of intake clinic and sMRI features for predicting language outcome of ASD toddlers.** Larger values of intake features with magenta color associated with better language outcome, while larger values of intake features with cyan color related to poorer language outcome.

**Table S1, Statistical results of ASD poor vs. TD and ASD good vs TD differences for brain regions showing significant GMV differences between ASD and TD.**

| **GMV Features** | **ASD Poor vs. TD** | | **ASD Good vs. TD** | |
|---|---|---|---|---|
| | **p** | **Cohen's d** | **p** | **Cohen's d** |
| Right Cerebellum Cortex | $3.76 \times 10^{-2}$ | -0.27 | $1.75 \times 10^{-2}$ | -0.33 |
| Posterior CC | $3.40 \times 10^{-4}$ | 0.46 | 0.60 | 0.07 |
| Mid Posterior CC | $7.37 \times 10^{-3}$ | 0.35 | 0.67 | 0.06 |
| Mid Anterior CC | $4.34 \times 10^{-4}$ | 0.46 | 0.86 | -0.02 |
| LH Fusiform | $1.08 \times 10^{-5}$ | 0.57 | 0.13 | 0.21 |
| LH Middle Temporal | $5.13 \times 10^{-4}$ | 0.45 | 0.06 | 0.27 |
| RH Caudal Anterior Cingulate | $4.39 \times 10^{-3}$ | -0.37 | $1.71 \times 10^{-3}$ | -0.44 |
| RH Middle Temporal | $1.41 \times 10^{-7}$ | 0.69 | 0.52 | 0.09 |

**Table S2, Statistical results of ASD poor vs. TD and ASD good vs TD differences for brain regions showing significant SA differences between ASD and TD.**

| SA Features | ASD Poor vs. TD | | ASD Good vs. TD | |
|---|---|---|---|---|
| | p | Cohen's d | p | Cohen's d |
| RH Caudal Anterior Cingulate | $2.85\times10^{-4}$ | -0.47 | $2.61\times10^{-4}$ | -0.51 |
| RH Medial Orbitofrontal | $1.01\times 10^{-6}$ | -0.64 | $3.93\times10^{-2}$ | -0.29 |
| RH Posterior Cingulate | $3.44\times 10^{-3}$ | -0.38 | $5.77\times10^{-4}$ | -0.48 |

**Table S3, Statistical results of ASD poor vs. TD and ASD good vs TD differences for brain regions showing significant thickness differences between ASD and TD.**

| Thickness Features | ASD Poor vs. TD | | ASD Good vs. TD | |
|---|---|---|---|---|
| | p | Cohen's d | p | Cohen's d |
| LH Caudal Middle Frontal | $7.68\times10^{-2}$ | -0.23 | $6.21\times10^{-4}$ | -0.48 |
| LH Pars Opercularis | $5.93\times10^{-3}$ | -0.36 | $1.19\times10^{-2}$ | -0.35 |
| LH Pericalcarine | $1.32\times10^{-2}$ | -0.32 | $2.89\times10^{-2}$ | -0.31 |
| LH Superior Temporal | $9.98\times10^{-4}$ | 0.43 | $5.17\times10^{-4}$ | 0.49 |
| RH Bank SSTS | $5.87\times10^{-3}$ | 0.36 | $3.87\times10^{-3}$ | 0.40 |
| RH Pars Opercularis | $2.09\times10^{-4}$ | -0.48 | 0.11 | -0.22 |
